# Reemergence of Oropouche virus between 2023 and 2024 in Brazil

**DOI:** 10.1101/2024.07.27.24310296

**Authors:** Gabriel C. Scachetti, Julia Forato, Ingra M. Claro, Xinyi Hua, Bárbara B. Salgado, Aline Vieira, Camila L. Simeoni, Aguyda R. C. Barbosa, Italo L. Rosa, Gabriela F. de Souza, Luana C. N. Fernandes, Ana Carla H. de Sena, Stephanne C. Oliveira, Carolina M. L. Singh, Shirlene T. de Lima, Ronaldo de Jesus, Mariana A. Costa, Rodrigo B. Kato, Josilene F. Rocha, Leandro C. Santos, Janete T. Rodrigues, Marielton P. Cunha, Ester C. Sabino, Nuno R. Faria, Scott C. Weaver, Camila M. Romano, Pritesh Lalwani, José Luiz Proença-Módena, William M. de Souza

**Affiliations:** Laboratory of Emerging Viruses, Department of Genetics, Evolution, Microbiology and Immunology, Institute of Biology, University of Campinas, Campinas, Brazil; Department of Microbiology, Immunology, and Molecular Genetics, College of Medicine, University of Kentucky, Lexington, KY, USA; Laboratory of Infectious Diseases and Immunology, Instituto Leônidas e Maria Deane, Fiocruz Amazônia, Manaus, Brazil; Universidade Federal do Amazonas, Manaus, Brazil; Fundação de Medicina Tropical Doutor Heitor Vieira Dourado, Manaus, Brazil; Laboratório Central de Saúde Pública do Ceará, Fortaleza, Brazil; Instituto de Ciências Biológicas, Universidade Federal de Minas Gerais, Belo Horizonte, Minas Gerais, Brazil; Laboratório Central de Saúde Pública do Acre, Rio Branco, Brazil; Instituto de Medicina Tropical, Faculdade de Medicina da Universidade de São Paulo, São Paulo, Brazil; Departamento de de Patologia, Faculdade de Medicina da Universidade de São Paulo, São Paulo, Brazil; MRC Centre for Global Infectious Disease Analysis, Department of Infectious Disease Epidemiology, School of Public Health, Imperial College London, London, UK; Department of Zoology, University of Oxford, Oxford, UK; Department of Microbiology and Immunology, University of Texas Medical Branch, Galveston, USA; World Reference Center for Emerging Viruses and Arboviruses, University of Texas Medical Branch, Galveston, USA; Hospital das Clínicas da Faculdade de Medicina da Universidade de São Paulo, São Paulo, Brazil

**Author notes:** These authors contributed equally to this work (Gabriel C. Scachetti, Julia Forato, Ingra M. Claro, Xinyi Hua, Bárbara B. Salgado, Marielton P. Cunha). These senior authors contributed equally to this work (Pritesh Lalwani, José Luiz Proença-Módena, William M. de Souza).

## Abstract

**Background:** Oropouche virus (OROV; species *Orthobunyavirus oropoucheense*) is an arthropod-borne virus that has caused outbreaks of Oropouche fever in Central and South America since the 1950s. This study investigates virological factors contributing to the reemergence of Oropouche fever in Brazil between 2023 and 2024.

**Methods:** In this study, we combined OROV genomic, molecular, and serological data from Brazil from 1 January 2015 to 29 June 2024, along with *in vitro* and *in vivo* characterization. Molecular screening data included 93 patients with febrile illness between January 2023 and February 2024 from the Amazonas State. Genomic data comprised two genomic OROV sequences from patients. Serological data were obtained from neutralizing antibody tests comparing the prototype OROV strain BeAn 19991 and the 2024 epidemic strain. Epidemiological data included aggregated cases reported to the Brazilian Ministry of Health from 1 January 2014 to 29 June 2024.

**Findings:** In 2024, autochthonous OROV infections were detected in previously non-endemic areas across all five Brazilian regions. Cases were reported in 19 of 27 federal units, with 83.2% (6,895 of 8,284) of infections in Northern Brazil and a nearly 200-fold increase in incidence compared to reported cases over the last decade. We detected OROV RNA in 10.8% (10 of 93) of patients with febrile illness between December 2023 and May 2024 in Amazonas. We demonstrate that the 2023-2024 epidemic was caused by a novel OROV reassortant that replicated approximately 100-fold higher titers in mammalian cells compared to the prototype strain. The 2023-2024 OROV reassortant displayed plaques earlier than the prototype, produced 1.7 times more plaques, and plaque sizes were 2.5 larger compared to the prototype. Furthermore, serum collected in 2016 from previously OROV-infected individuals showed at least a 32-fold reduction in neutralizing capacity against the reassortment strain compared to the prototype.

**Interpretation:** These findings provide a comprehensive assessment of Oropouche fever in Brazil and contribute to a better understanding of the 2023-2024 OROV reemergence. The recent increased incidence may be related to a higher replication efficiency of a new reassortant virus that also evades previous immunity.

## Introduction

Oropouche virus (OROV) is an endemic and neglected arbovirus that causes Oropouche fever in Central and South America^1,2^. Oropouche fever is usually characterized by mild and self-limited, non-specific disease with headache, arthralgia, myalgia, nausea, vomiting, chills, and photophobia^1^. However, some patients can experience more severe complications, such as hemorrhagic manifestations, meningitis, or meningoencephalitis^3,4^. OROV is probably transmitted in the enzootic cycle mainly by *Culicoides paraensis* midges among pale-throated sloths (*Bradypus tridactylus*), non-human primates and other wild mammals^1,5^. In urban areas, the human-amplified cycle appears to have been established on multiple occasions involving midges and humans, and potentially some mosquito species^1,5^. Neither licensed vaccines nor antiviral drugs are available to prevent OROV infection or to treat Oropouche fever.

OROV belongs to the species *Orthobunyavirus oropoucheense* in the *Orthobunyavirus* genus of the *Peribunyaviridae* family, and its genome consists of three single-stranded, negative-sense RNA molecules: small (S), medium (M), and large (L) segments^6^. Like other multi-segmented viruses, OROV can undergo reassortment events when two related orthobunyaviruses co-infect the same cell, resulting in progeny with mixed genomic segments from both parental strains^7^. These events are important drivers of genetic divergence because they can alter vector competence and disease severity^8^. For instance, three OROV reassortants have been identified in South America: Iquitos (IQTV), Madre de Dios, and Perdões viruses^2^.

OROV infections have been documented in Central and South America since the 1950s^1,2^. Although the burden of Oropouche fever remains unknown, some estimates suggest over half a million human cases may have occurred since its first identification^1^. Between November 2023 and June 2024, a substantial increase in Oropouche fever cases was observed in Brazil, Bolivia, Colombia, and Peru, and autochthonous cases were detected in Cuba^9^. Here, we contextualize the Oropouche fever spread in Brazil from 2015 to 2024 and combine epidemiological, molecular, genomic, and serological analysis to describe and investigate the virological factors that may have contributed to Oropouche fever reemergence.

## Methods

### Study design and setting

This study combined multiple data sources from OROV including molecular, genomic, and serological data with aggregated epidemiological data from Brazil between 1 January 2015 and 29 June 2024. National epidemiological data of laboratory-confirmed cases (e.g., RT-qPCR and ELISA) were obtained from the Brazilian Ministry of Health. This dataset includes the aggregated number of Oropouche fever cases in Brazil from epidemiological week 1 (04 to 10 January) in 2015 to epidemiological week 26 in 2024 (23 to 29 June) and Oropouche fever cases per state. Additionally, for molecular screening, we used residual serum samples from 93 patients with acute febrile illnesses negative for malaria collected between 15 January and 5 February 2024. All serum samples were obtained from patients cared for in the public health system, for whom samples were submitted to the Fundação de Medicina Tropical Doutor Heitor Vieira Dourado in Manaus City, as part of the surveillance system of Amazonas State, Brazil. All procedures followed the ethical standards of the responsible committee on human experimentation and were approved by the ethics committees from the Federal University of Amazonas, Brazil (approval numbers 5.876.612 and 6.629.451) and Nilton Lins University (2.636.421).

### Procedures

For the patients sampled in our molecular screening, basic clinical and demographic data were collected from the Brazilian Laboratorial Environment Management System. Anonymized patient information data were used in the current study and are provided in the **appendix, Table S1**. Viral RNA was extracted from serum samples and tested for OROV RNA using a specific real-time quantitative Reverse-Transcription Polymerase Chain Reaction (RT-qPCR) assay^10,11^. RT-qPCR-positive RNA samples were used for viral isolation in Vero CCL81 cells. Isolated viruses were confirmed by RT-qPCR and focus-forming assay (FFA) (**appendix, p 1-2**). Viral isolates were sequenced using the nanopore platform and subjected to phylogenetic analysis (**appendix, p 2**). Next, to assess susceptibility to antibody neutralization, the 2023-2024 epidemic isolate and OROV prototype strain BeAn 19991 (prototype) were compared using sera from 22 individuals with confirmed OROV infection from Coari municipality, Amazonas State, Brazil. All these convalescent sera exhibited high neutralizing titers measured by a plaque reduction neutralization test (PRNT_50_) (**appendix, p 1-3**). Additionally, to evaluate cross-neutralization between the OROV 2023-2024 epidemic and BeAn 19991 isolates, we inoculated four-week-old C57BL/6 mice intraperitoneally. After four weeks, blood was collected, and a PRNT_50_ was performed to assess antigenic differences between the viral strains (**appendix, p 3**). All animal procedures from this study were approved by the Ethics Committee on Animal Use of the University of Campinas (approval numbers 5171-1/2019, 6003-1/2022, and 5657-1/2020).

### Statistical analysis

The analyses were carried out in RStudio version 4.4.0 (https://rstudio.com). Incidences were calculated based on the 2022 Brazilian population census reported by the Brazilian Institute of Geography and Statistics (www.ibge.gov.br). Statistical analyses of the viral replication curves, the number of plaques, and plaque size were performed using the paired t-test, as indicated in the figure legends. To assess whether serum PRNT_50_ against isolate of the 2023-2024 epidemic were reduced compared with those against OROV strain BeAn 19991 in individuals with a previous OROV infection, we used a binomial distribution model based on the proportion of samples in each of two categories (differences >0 and differences ≤0) as previously described^12^. For analysis of differences in plasma neutralizing antibody titers, median PRNT_50_ for each sample was calculated as a mean of two technical duplicates, and statistical significance of the differences between group medians was determined by one-way ANOVA followed by Tukey’s test of non-linear regression curves using GraphPad Prism software version 8.2.1.

## Results

Between January 1, 2015, and June 29, 2024, 8,284 laboratory-confirmed Oropouche fever cases were reported to the Brazilian Ministry of Health. Of these, 84.2% (6,973 of 8,284) were reported from January to June 2024, representing 199 times more than the annual median of 35 cases (interquartile, 23-91) reported between 2015 and 2023 (**Fig. 1a**). Oropouche cases peaked annually between November and March, with 88.7% (7,253 of 8,176) of all cases reported during these months. Northern Brazil was the most affected region, with 83.2% (6,895 of 8,284) of all cases reported between 2015 and 2024 (**Fig. 1b**). Amazonas state reported the highest number of cases (44.7%, 3,699 of 8,284) and a cumulative incidence of 93.8 cases per 100,000 inhabitants from 2015 to 2024. Prior to 2024, Oropouche fever cases in Brazil primarily affected Northern Brazil. However, the 2024 epidemic resulted in widespread disease across Brazil, with autochthonous cases being identified in all five Brazilian regions. These newly affected regions included the Northeast (Bahia, Ceará, Pernambuco, and Piauí States), the Southeast (Espírito Santo, Minas Gerais, and Rio de Janeiro States), Central-west (Mato Grosso and Mato Grosso do Sul States) and the South (Santa Catarina State). The incidence of the disease in newly affected areas ranged from 0.03 to 9.8 cases per 100,000 inhabitants (**Fig. 1b**). Our analysis of the age-sex structure of Oropouche fever cases between 2015 and 2024 revealed that individuals of both sexes aged 20 to 49 years exhibited a higher incidence compared to the overall average incidence of 4.1 cases per 100,000 inhabitants (**Fig. 1c**).

**Figure 1.**
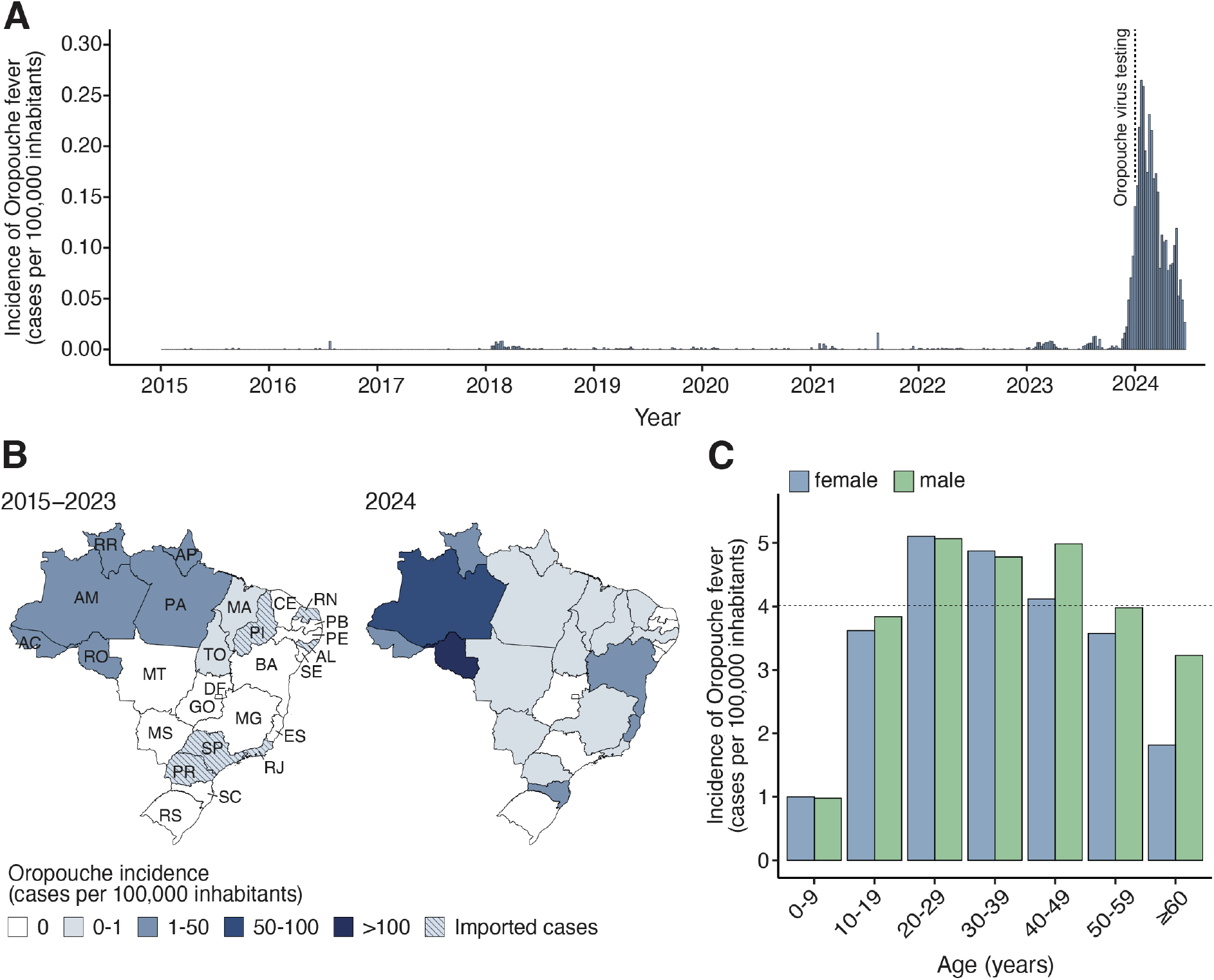
Spatial-temporal dynamics of Oropouche fever between 2015 and 2024 in Brazil. **(A)** Incidence of laboratory-confirmed Oropouche fever cases per epidemiological week in all 26 Brazilian States and the Federal District, from epidemiological week 1 of 2015 (04 to 10 January) to epidemiological week 26 of 2024 (23 to 29 June). The dashed line indicates the implementation of OROV diagnosis across all Central Public Health Laboratories in Brazil. **(B)** Maps were colored according to the incidence of laboratory-confirmed Oropouche fever cases by federal unit between January 2015 and December 2023 (left) and January to June 2024 (right). AC=Acre. AL=Alagoas. AM=Amazonas. AP=Amapá. BA=Bahia. CE=Ceará. ES=Espírito Santo. DF=Distrito Federal (Federal District). GO=Goiás. MA=Maranhão. MG=Minas Gerais. MS=Mato Grosso do Sul. MT=Mato Grosso. PA=Pará. PB=Paraíba. PE=Pernambuco. PI=Piauí. PR=Paraná. RJ=Rio de Janeiro. RN=Rio Grande do Norte. RO=Rondônia. RR=Roraima. RS=Rio Grande do Sul. SC=Santa Catarina. SE=Sergipe. SP=São Paulo. TO=Tocantins. **(C)** Oropouche fever incidence based on the age-sex distribution of cases from 2015 to 2024.

We next used RT-qPCR to investigate active OROV circulation in 93 patients with acute febrile illness from December 2023 to March 2024 in Amazonas State. OROV RNA was detected in 10.8% (10 of 93) of patients with acute febrile illness, with viral loads ranging from 1.65×10^3^ to 3.4×10^4^ PFU/mL as determined using a standard curve of infectious viruses titrated by FFA. We also detected DENV RNA in two patients infected with serotypes 1 and 2. Chikungunya, Mayaro (MAYV), and dengue viruses (DENV) serotypes 3 and 4 were not detected. We isolated OROV in Vero CCL-81 cells, with cytopathic effects (CPE) observed within approximately 30 hours post-inoculation (**Fig. 2a**). Three blind passages were performed, and viral RNA was confirmed in 70% (7 of 10) of passaged strains using RT-qPCR on culture cell passages exhibiting CPE. Next, we assessed viral titers using FFA in all three passages and compared them with the original samples, showing the increased viral loads of passaged OROV isolates compared to the original patient samples (**Fig. 2b**).

**Figure 2.**
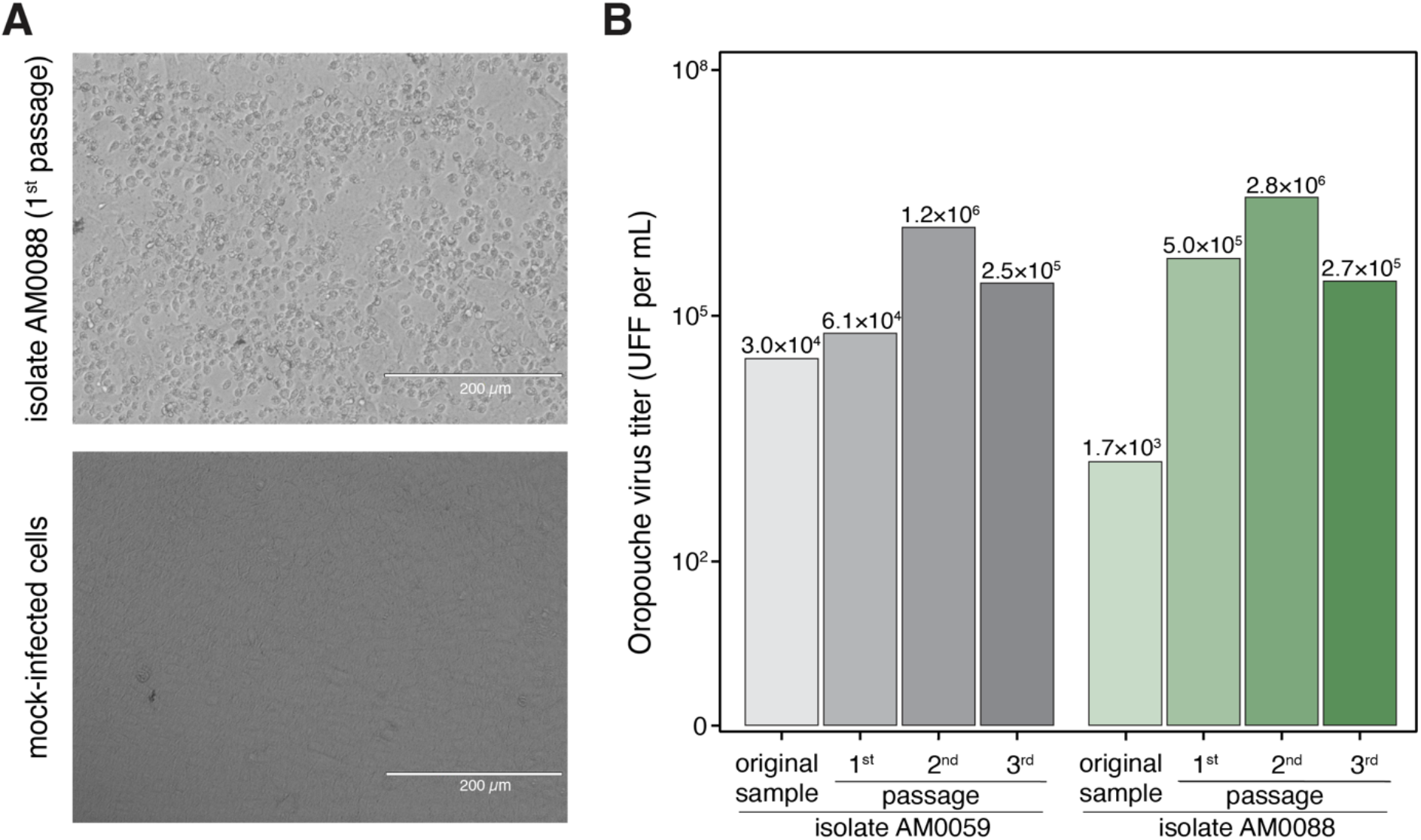
Oropouche virus from the 2024 epidemic (AM0059 and AM0088) isolated from serum samples of Oropouche fever patients from Manaus City, Amazonas, Brazil. **(A)** Isolation of the 2024 epidemic of OROV was performed in Vero CCL81 cells, and a typical cytopathic effect was observed approximately 30 hours post-infection in inoculated cells in comparison with uninfected cells. (**B**) The isolation was also confirmed by focus forming assay, where it is possible to observe an increase in viral load. Images were obtained in an EVOS inverted microscope kindly provided by Thermo Fisher Scientific.

We subsequently sequenced the nearly complete coding sequences of two OROV isolates (i.e., AM0059 and AM0088). We obtained over 90% coverage of two OROV genomes (all three segments) with a mean depth of coverage of ≥ 20-fold per nucleotide. We submitted sequences to GenBank (accession nos. PP992525 – PP992530). Maximum likelihood phylogenetic analyses showed that the AM0059 and AM0088 strains clustered within a highly supported monophyletic clade (bootstrap support 100%) within 2022-2023 strains Amazonas, Acre, Roraima, and Rondônia States (**Fig. 3**). Based on the topology of phylogenetic trees estimated for the three OROV segments and the RDP4 analysis with concatenated genomes, we found that 2023-2024 OROV strains resulted from a reassortment event, as previously described^13^. The L protein (L segment) of AM0059 and AM0088 shared 95-96% amino acid identity with the OROV strain BeAn 19991, which represents 65 amino acid differences between them. The glycoprotein (segment M) shared 98% identity with up to 31 amino acid differences (**appendix, Table S2**). No amino acid changes were found in the nucleoprotein (segment S) of AM0059 and AM0088 compared to BeAn 19991 strain. During molecular screening for OROV using two RT-qPCR protocols^10,11^, we observed that samples from 10 patients tested positive for one protocol but were all negative using RT-qPCR other protocol targeting segment S^11^. We found that mismatches between the segment S of 2023-2024 OROV reassortant strains and the primers and probes likely impacted these diagnoses (**appendix, Fig. S1**).

**Figure 3.**
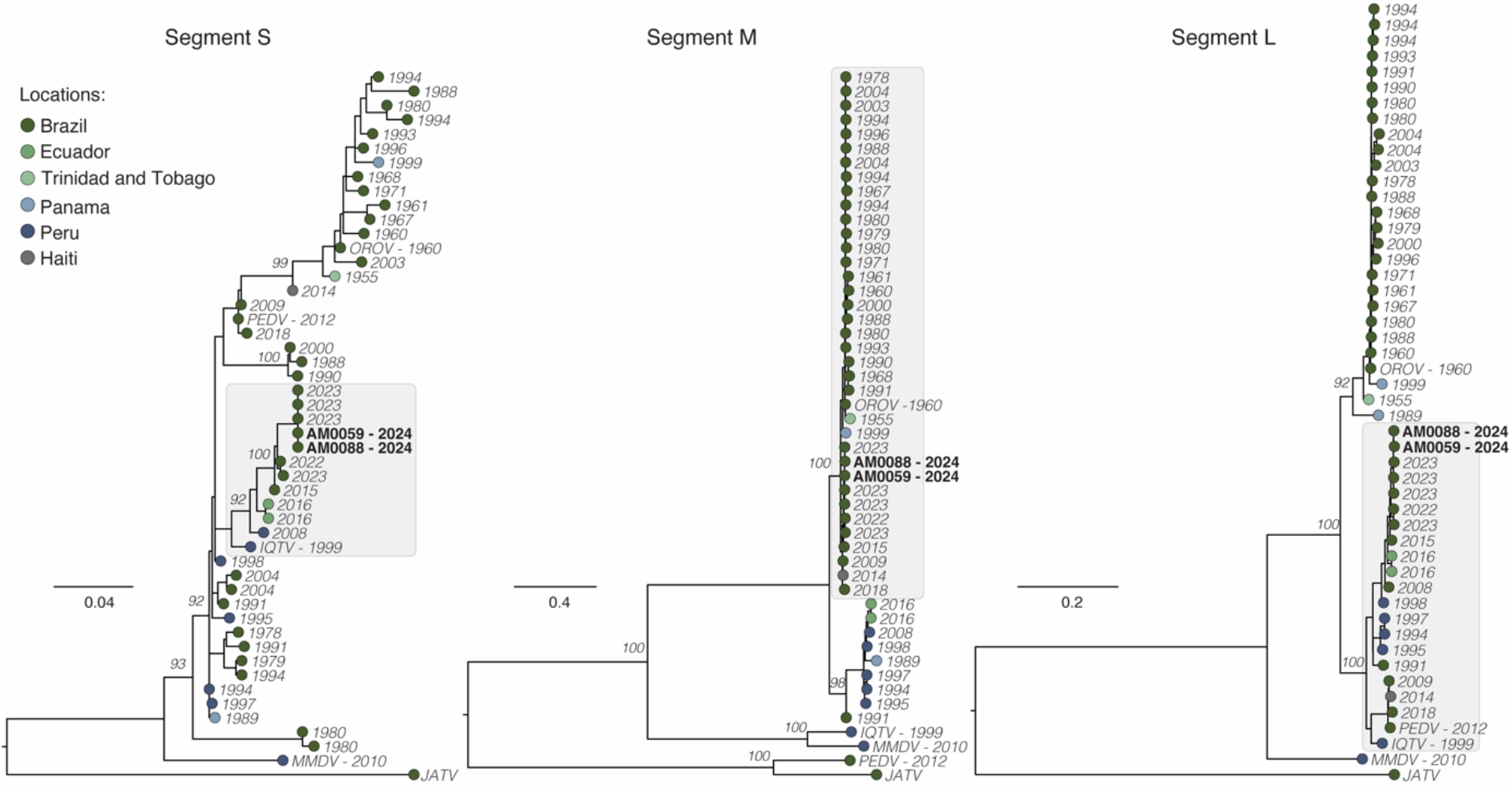
Phylogenetic analysis of the Oropouche virus. Maximum likelihood phylogenetic tree of 48 representative OROV genomes, including two new genomes (black bold) from Manaus City generated in this study. Phylogenetic trees are shown for the S segment (left), M segment (center), and L segment (right). Tips are colored according to the location country of each sample. Phylogenies were midpoint rooted for clarity of presentation. Scale bar indicates the evolutionary distance of substitutions per nucleotide site. Bootstrap values based on 1,000 replicates are shown on principal nodes. The GenBank accession numbers of sequences used in this figure are presented in Appendix Table S4.

Next, we compared the replication kinetics of the 2024 epidemic isolate (OROV strain AM0088) and OROV strain BeAn 19991 in Vero CCL81, Huh7, and U-251 cell lines. Cells were infected at a multiplicity of infection (MOI) of 0.1, and infectious virus was quantified by FFA at 3-, 6-, 12-, and 24-hours post-infection (hpi). We found a significantly higher replication of the AM0088 strain compared to BeAn 19991 in all cell lines at 12 and 24 hpi, with a median difference of 157 (interquartile, 57-394) times at these specified time points (**Fig. 4a**). We found that the AM0088 strain formed plaques at 36 hpi, while BeAn 19991 presented plaques at 48 hpi. Additionally, the AM0088 strain formed 66% and 27% more plaques at 48 and 72 hpi compared to BeAn 19991 (**Fig. 4b**). The AM0088 strain also exhibited a distinctly larger plaque phenotype compared to BeAn 19991, with 2.6-fold and 2.4-fold larger diameters at 48 and 72 hpi (**Fig. 4c**).

**Figure 4.**
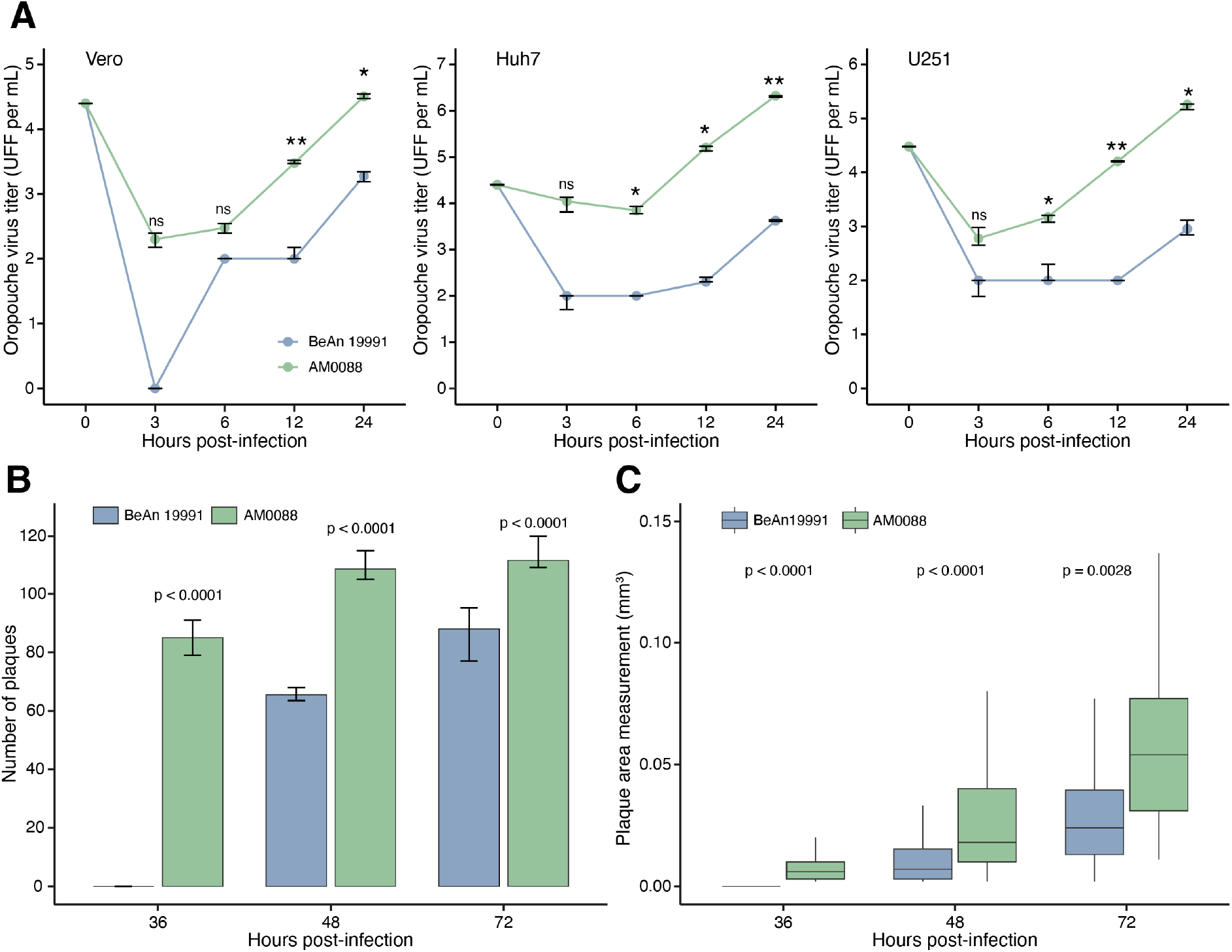
Characterization in vitro of 2024 Oropouche virus reassortment strain. **(A)** Viral replication properties of OROV strain AM0088 and OROV strain BeAn 19991 in Vero CCL81 (left), Huh7 (center), and U-251 (right) cell lines. Cells were infected at a multiplicity of infection of 0.1. At indicated time points, samples were harvested, and titers were determined by focus-forming assay on Vero CCL81 cells. Statistical analyses were performed using the paired t-test. The median is presented by the middle line, and the upper and lower limits represent the 75^th^ and 25^th^ percentiles (whiskers). Statistical significance is ***p < 0.001, **p < 0.01, and *p < 0.05; ns, not significant. **(B)** Number of plaques formation by OROV strains AM0088 and BeAn 19991 at 36-, 48-, and 72-hpi in Vero CCL8 cells (n=12 wells). **(C)** Size of plaques by OROV strains AM0088 and BeAn 19991 at 36-, 48-, and 72-hpi in Vero CCL81 cells (n≥ 118 plaques).

Subsequently, we investigate whether the AM0088 strain might escape from neutralization by antibodies induced by previous OROV infection. For this analysis, we used serum samples collected in May 2016 from 22 individuals previously infected with OROV in Coari municipality, Amazonas State (**appendix 2, p 1**). Using a PRNT_50_ assay, we assessed the ability of these serum samples to neutralize both the BeAn 19991 and the AM0088 isolates. We found that serum from individuals previously infected with OROV had a median PRNT_50_ for BeAn 19991 of 640 (interquartile 320 – 640), while the same serum had PRNT_50_ titers below the limit of detection (<20) against AM0088 isolate (p < 0.0001) (**Fig. 5a**). These findings suggest a roughly minimum 32-fold reduction in the neutralizing capacity of antibodies from previously infected individuals to target the AM0088 compared to the BeAn 19991 isolate. Lastly, we conducted cross-neutralization tests to explore the serological relationships between AM0088 and BeAn 19991 isolates after experimental infection of C57BL/6 mice. The serum samples collected 28 days after mice were infected with the AM0088 strain showed a mean homologous neutralizing antibodies titer of 320 (interquartile 160 – 320) against the AM0088 strain and a titer of ≥ 640 against BeAn 19991 (**Fig. 5b**). Additionally, the serum samples from mice immunized with BeAn 19991 strain show a neutralizing antibodies titer of ≥ 640 against the BeAn 19991 strain and a titer of 80 (interquartile 40 – 80) for the AM0088 strain (**Fig. 5c**). This indicates that AM0088 reassortant is serologically distinct and significantly less neutralized by antibodies generated through infection with BeAn 19991.

**Figure 5.**
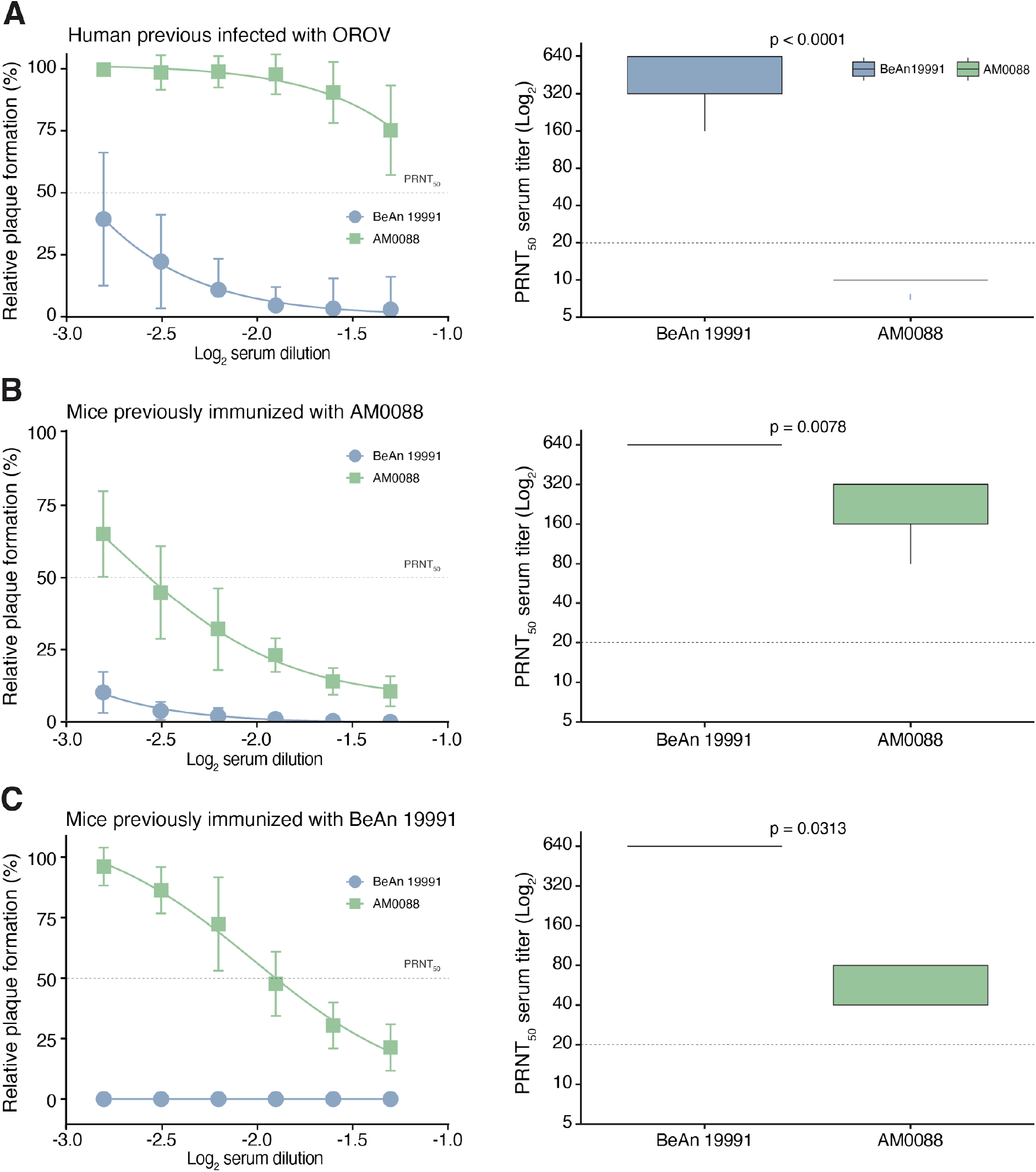
Neutralization of Oropouche virus strains BeAn 19991 and AM0088 by PRNT_50_. **(A)** Serum samples were tested by PRNT_50_ in Vero CCL81 cells after incubation with OROV strain BeAn 19991 or AM0088. PRNT_50_ represents the sample dilution that showed a 50% reduction in plaque formation compared with a control well inoculated with OROV alone (without serum), after linear regression analysis. Each data point represents the mean of all serum samples for each group at each dilution level (shown as log_2_ serum dilution), and error bars represent SD (left). PRNT_50_ titer that is defined as the reciprocal value of the serum dilution that reduced cytopathic effects by 50% against cytopathic effects (right). **(A)**. Serum samples individuals previously infected with OROV in Coari, Amazonas State, Brazil (n=22). **(B)**. Serum samples were collected from C57BL/6 mice 28 days after infection with OROV strain AM0088 (n=7). **(C)**. Serum samples were collected from C57BL/6 mice 28 days after infection with OROV strain BeAn 19991 (n=5).

## Discussion

This study provides a comprehensive assessment of Oropouche fever in Brazil from 2015 to 2024, focusing on the 2023-2024 reemergence, which had an approximately 200-fold higher incidence (laboratory-confirmed cases) than between 2015 and 2023. The substantial increase in the number of cases can be partially attributed to the increase of OROV surveillance in public health laboratories across Brazil^14^. We found that Amazonas was the most affected Brazilian state, where the reemergence in 2023-2024 was caused by a new OROV reassortant. This reassortant involved the M segment from a previously circulating OROV strain in Northern Brazil combined with S and L segments from a previous reassortant originated from IQTV, as previously described^13^. Our data suggest that high neutralizing antibodies from prior OROV infection may not efficiently neutralize with this new OROV reassortment. Similarly, a previous study using convalescent human sera and mouse antisera demonstrated that prior OROV infection does not provide protection against IQTV, a reassortant identified in Peru in 1999^15^. These findings collectively highlight the limited protective immunity against OROV and its reassortants in Brazil, suggesting individuals may be reinfected and develop clinical disease.

Our findings show that the 2023-2024 OROV reassortant exhibits significantly higher replicative competence in mammalian cells (i.e., Vero CCL81, Huh7, and U-251) compared to the ancient OROV strain BeAn 19991. The extent to which this faster replication translates to enhanced OROV transmissibility by vectors remains unclear, but it is plausible that this viral fitness may lead to an increase in viremia, resulting in more efficient infection of vectors during blood feeding^16,17^. Another common measure of viral fitness is the number and size of plaques, which are caused by necrosis or apoptosis upon infection of a cell monolayer^18-20^. Our data show that the genotypic changes induced by the 2023-2024 OROV reassortment mediate an increase in the number and size of plaques, which suggests increased virulence compared to the prototype strain^19,20^. Therefore, this phenotypic change might partially explain the observed increase in OROV incidence between November 2023 and June 2024, but further investigation is necessary to elucidate the molecular mechanisms underlying this altered plaque morphology in the new OROV reassortment.

Our analysis revealed a higher incidence of Oropouche fever in 2024 within Northern Brazil, a historically endemic region with documented OROV circulation since the 1950s^1^. This observation suggests that human populations in these areas continue to be exposed to OROV and might be susceptible to reinfection due to reduced neutralizing antibody capacity against new epidemic OROV variants, such as the 2023-2024 OROV reassortment. This hypothesis is based on the principle that prior OROV infection elicits a robust humoral response that can protect against homologous infection but not against novel reassortments, as previously described for IQTV^21^. This is because the main target of the neutralizing antibody response for orthobunyaviruses is the trimeric spikes in the glycoprotein encoded by the M segment^22^, and consequently, change of this segment or mutations in glycoprotein gene may lead to reduced neutralizing antibody binding. Furthermore, the 2023-2024 surveillance data reveals a geographical expansion of Oropouche fever into previously non-endemic areas, including more densely populated states in Brazil, such as Bahia, Piauí, Ceará, Espírito Santo, Minas Gerais, Rio de Janeiro, and Santa Catarina States. This spread highlights the risk posed by the large immunologically naïve population across the Americas combined with the widespread availability of *C. paraensis* and other potential OROV vectors from the southeastern United States to Uruguay^23^. Consequently, the reduced neutralizing antibody levels and putative increased viral fitness of the novel reassortment may contribute to reinfections in endemic areas like the Amazon Basin but also increase spread into new regions. This scenario could contribute to the introduction and establishment of OROV in the Americas and beyond, such as previously observed with DENV, ZIKV, and CHIKV^24^.

We found that epidemic peaks of OROV cases in the epidemic waves occur mainly between November and March, which coincides with the rainy season and higher temperatures in the Brazilian Amazon region. Wet and warm periods have been described as critical drivers of the magnitude and seasonality of vector-borne virus transmission based on how they affect vector reproduction, survival, biting rates, and population density^25^. Previous studies show that *Culicoides paraensis* has the greatest abundance in the rainy season, with temperatures between 30°C and 32°C and relative air humidity between 75% and 85%, with a substantial decline during the dry season^26^. However, the lack of information about the impact of climate and weather on *C. paraenesis* and other potential OROV vectors prevents exploring the potential impact of higher temperatures and the 2023-2024 El Niño event as potential ecological factor associated with the OROV reemergence. Additionally, our data indicate that OROV affected predominantly adults aged 20 to 49 years old from both sexes. However, a comprehensive understanding of risk factors requires further exploration of ecological and individual-level determinants, including socioeconomic status, occupation, and deprivation^27^.

This study presents several limitations. First, the absence of historical data from the convalescent OROV-infection individuals prevents us from determining the OROV strain involved and estimating the time since previous exposure. However, the higher neutralizing antibody titers against BeAn 19991 suggest that previous infections in these individuals were likely caused by a similar OROV strain. Second, neutralizing antibodies against orthobunyaviruses primarily target the glycoproteins encoded by the M segment. The S and L segments of the 2023-2024 OROV reassortment originated from IQTV, but the M segment contains 31 amino acid substitutions compared to BeAn1991. Therefore, it is plausible that these amino acid changes may have a more significant impact on viral fitness in humans and vectors than the reassortment itself. Further studies using reverse genetics techniques are necessary to elucidate the relative contributions of these factors. Third, our study focused on neutralizing antibodies, but cellular immune responses mediated by T cells or functions mediated by Fc-mediated antibody effector response may also influence protection against disease^28,29^. Additional studies are required to clarify the protective role of prior OROV exposure against reinfection and determine the duration of protective immunity. Another limitation is that our study relied on variable healthcare-seeking behaviors, and the previous lack of molecular diagnostic capabilities for OROV at national level may have contributed to underestimating the Oropouche fever burden in Brazil in the previous years. Likewise, we were unable to determine the full extent of mild or asymptomatic cases due to the challenges of syndromic surveillance in regions co-endemic for DENV, MAYV, and CHIKV, all of which cause similar clinical presentations to Oropouche fever^24,30^.

In conclusion, our findings provide an important context about the dynamics and drivers of OROV and may inform future studies and public health policy focusing on the strategies to mitigate the impact of new outbreaks and epidemics. For example, the 2023-2024 OROV reemergence underscores the need for robust molecular surveillance strategies targeting at least two viral genome segments. Also, OROV infection should be considered in the differential diagnosis of febrile illnesses and neurological complications. Further investigation is warranted to define the burden of severe Oropouche fever disease. These findings emphasize the need for public health preparedness strategies and the development of vaccines that provide broader protection against OROV and its reassortments to respond to future outbreaks effectively.

## Data Availability

All statistical computing analyses were conducted using the R project. R packages necessary for analysis and visualization include: readxl, tidyverse, dylyr, zoo, scales, lubridate, tmap, sf, ggplot2, ggbreak, ggpubr, stringr. No custom code was developed. New sequences have been deposited in GenBank with accession numbers PP992525 - PP992530.

## Acknowledgments

JLPM is supported by São Paulo Research Foundation (#2022/10442-0), and National Council for Scientific and Technological Development (CNPq, #309971/2023-3). WMdS is supported by Burroughs Wellcome Fund – Climate Change and Human Health Seed Grants (#1022448), and Wellcome Trust – Digital Technology Development Award in Climate Sensitive Infectious Disease Modelling (#226075/Z/22/Z). NRF was supported by a Wellcome Trust and Royal Society Sir Henry Dale Fellowship (grant no. 204311/Z/16/Z). NRF and ECS are supported by a Medical Research Council-São Paulo Research Foundation - CADDE partnership award (MR/S0195/1 and FAPESP 18/14389-0). SCW was supported by NIH grant R24 AI120942. CMR is supported by São Paulo Research Foundation (#2022/10408-6). IMC was supported by São Paulo Research Foundation (#2023/11521-3). CLS was supported by São Paulo Research Foundation (#2022/00723-1). PL is supported by Fundação de Amparo a Pesquisa do Estado do Amazonas (POSGRAD, CT&I ÁREAS PRIORITÁRIAS, INICIATIVA AMAZÔNIA +10, CHAMADA ERC 2022 and PEX-CT&I/FAPEAM); Coordenação de Aperfeiçoamento de Pessoal de Nível Superior, Funding 001). We thank Jacqueline A. Tida (www.plotmyscience.com) for the figure editing.

## Contributions

GCS, JF, PL, JLPM, and WMdS conceptualized the study. JF, GCS, IMC, BBS, GFdS, ARCB. ILR, LCNF, ACHdS, SCP, CMLS, STdL, CLS, AV, MAC, JFR, LCS, JTR, MPC, ECS, CMR, PL, JLMP, and WMdS contributed to the acquisition of data. JF, GCS, IMC, XH, RdJ, RBK, JLMP, and WMdS contributed to the data analysis. JF, GCS, IMC, XH, NRF, SCW, PL, JLMP, and WMdS contributed to data interpretation. JF, GCS, JLMP, and WMdS drafted the manuscript. JF, GCS, ECS, CMR, SCW, NRF, PL, JLMP, and WMdS revised the manuscript. Funding: ECS, CMR, PL, JLMP, and WMdS acquired funding for the study. All authors read and approved the final version of the manuscript and had access to all the data in the study. JLMP and WMdS accessed and verified all the data reported in the study.

## Data sharing

All statistical computing analyses were conducted using the R project. R packages necessary for analysis and visualization include: readxl, tidyverse, dylyr, zoo, scales, lubridate, tmap, sf, ggplot2, ggbreak, ggpubr, stringr. No custom code was developed. New sequences have been deposited in GenBank with accession numbers PP992525 – PP992530.

## References

1. Travassos da Rosa JF, de Souza WM, Pinheiro FP, et al. Oropouche Virus: Clinical, Epidemiological, and Molecular Aspects of a Neglected Orthobunyavirus. Am J Trop Med Hyg 2017; 96(5): 1019–30.

2. Wesselmann KM, Postigo-Hidalgo I, Pezzi L, et al. Emergence of Oropouche fever in Latin America: a narrative review. Lancet Infect Dis 2024.

3. Bastos MeS, Figueiredo LT, Naveca FG, et al. Identification of Oropouche Orthobunyavirus in the cerebrospinal fluid of three patients in the Amazonas, Brazil. Am J Trop Med Hyg 2012; 86(4): 732–5.

4. Vernal S, Martini CCR, da Fonseca BAL. Oropouche Virus-Associated Aseptic Meningoencephalitis, Southeastern Brazil. Emerg Infect Dis 2019; 25(2): 380–2.

5. Pinheiro FP, Travassos da Rosa AP, Gomes ML, LeDuc JW, Hoch AL. Transmission of Oropouche virus from man to hamster by the midge Culicoides paraensis. Science 1982; 215(4537): 1251–3.

6. Hughes HR, Adkins S, Alkhovskiy S, et al. ICTV Virus Taxonomy Profile:. J Gen Virol 2020; 101(1): 1–2.

7. Elliott RM. Orthobunyaviruses: recent genetic and structural insights. Nat Rev Microbiol 2014; 12(10): 673–85.

8. Briese T, Bird B, Kapoor V, Nichol ST, Lipkin WI. Batai and Ngari viruses: M segment reassortment and association with severe febrile disease outbreaks in East Africa. J Virol 2006; 80(11): 5627–30.

9. Moutinho S. Little-known virus is on the rise in South America. Science 2024; 384(6700): 1052–3.

10. Naveca FG, Nascimento VAD, Souza VC, Nunes BTD, Rodrigues DSG, Vasconcelos PFDC. Multiplexed reverse transcription real-time polymerase chain reaction for simultaneous detection of Mayaro, Oropouche, and Oropouche-like viruses. Mem Inst Oswaldo Cruz 2017; 112(7): 510–3.

11. de Souza Luna LK, Rodrigues AH, Santos RI, et al. Oropouche virus is detected in peripheral blood leukocytes from patients. J Med Virol 2017; 89(6): 1108–11.

12. Souza WM, Amorim MR, Sesti-Costa R, et al. Neutralisation of SARS-CoV-2 lineage P.1 by antibodies elicited through natural SARS-CoV-2 infection or vaccination with an inactivated SARS-CoV-2 vaccine: an immunological study. Lancet Microbe 2021.

13. Naveca FG, Almeida TAP, Souza VS, et al. Emergence of a novel reassortant Oropouche virus drives persistent outbreaks in the Brazilian Amazon region from 2022 to 2024. Virological. https://virological.org/t/emergence-of-a-novel-reassortant-oropouche-virus-drives-persistent-outbreaks-in-the-brazilian-amazon-region-from-2022-to-2024/955; 2024.

14. Ministério da Saúde do Brasil. Orientações para a vigilância da Febre do Oropouche. NOTA TÉCNICA Nº 6/2024-CGARB/DEDT/SVSA/MS. In: Departamento de Doenças Transmissíveis Coordenação-Geral de Vigilância de Arboviroses. https://www.gov.br/saude/pt-br/centrais-de-conteudo/publicacoes/notas-tecnicas/2024/nota-tecnica-no-6-2024-cgarb-dedt-svsa-ms; 2024.

15. Forshey BM, Guevara C, Laguna-Torres VA, et al. Arboviral etiologies of acute febrile illnesses in Western South America, 2000–2007. PLoS Negl Trop Dis 2010; 4(8): e787.

16. Duong V, Lambrechts L, Paul RE, et al. Asymptomatic humans transmit dengue virus to mosquitoes. Proc Natl Acad Sci U S A 2015; 112(47): 14688–93.

17. Rückert C, Ebel GD. How Do Virus-Mosquito Interactions Lead to Viral Emergence? Trends Parasitol 2018; 34(4): 310–21.

18. Goh KC, Tang CK, Norton DC, et al. Molecular determinants of plaque size as an indicator of dengue virus attenuation. Sci Rep 2016; 6: 26100.

19. Wargo AR, Kurath G. Viral fitness: definitions, measurement, and current insights. Curr Opin Virol 2012; 2(5): 538–45.

20. Domingo E, de Ávila AI, Gallego I, Sheldon J, Perales C. Viral fitness: history and relevance for viral pathogenesis and antiviral interventions. Pathog Dis 2019; 77(2).

21. Aguilar PV, Barrett AD, Saeed MF, et al. Iquitos virus: a novel reassortant Orthobunyavirus associated with human illness in Peru. PLoS Negl Trop Dis 2011; 5(9): e1315.

22. Hellert J, Aebischer A, Wernike K, et al. Orthobunyavirus spike architecture and recognition by neutralizing antibodies. Nat Commun 2019; 10(1): 879.

23. Files MA, Hansen CA, Herrera VC, et al. Baseline mapping of Oropouche virology, epidemiology, therapeutics, and vaccine research and development. NPJ Vaccines 2022; 7(1): 38.

24. de Souza WM, Ribeiro GS, de Lima STS, et al. Chikungunya: a decade of burden in the Americas. Lancet Reg Health Am 2024; 30: 100673.

25. Walsh JF, Molyneux DH, Birley MH. Deforestation: effects on vector-borne disease. Parasitology 1993; 106 Suppl: S55–75.

26. Feitoza LHM, de Carvalho LPC, da Silva LR, et al. Influence of meteorological and seasonal parameters on the activity of Culicoides paraensis (Diptera: Ceratopogonidae), an annoying anthropophilic biting midge and putative vector of Oropouche Virus in Rondônia, Brazilian Amazon. Acta Trop 2023; 243: 106928.

27. Power GM, Vaughan AM, Qiao L, et al. Socioeconomic risk markers of arthropod-borne virus (arbovirus) infections: a systematic literature review and meta-analysis. BMJ Glob Health 2022; 7(4).

28. Ribeiro Amorim M, Cornejo Pontelli M, Fabiano de Souza G, et al. Oropouche Virus Infects, Persists and Induces IFN Response in Human Peripheral Blood Mononuclear Cells as Identified by RNA PrimeFlow™ and qRT-PCR Assays. Viruses 2020; 12(7).

29. Proenca-Modena JL, Sesti-Costa R, Pinto AK, et al. Oropouche virus infection and pathogenesis are restricted by MAVS, IRF-3, IRF-7, and type I interferon signaling pathways in nonmyeloid cells. J Virol 2015; 89(9): 4720–37.

30. Forato J, Meira CA, Claro IM, et al. Molecular Epidemiology of Mayaro Virus among Febrile Patients, Roraima State, Brazil, 2018-2021. Emerg Infect Dis 2024; 30(5): 1013–6.

