## Appendix for "Reemergence of Oropouche virus between 2023 and 2024 in Brazil"

**Supplementary appendix**

**Supplementary Information: Materials and Methods**

**Serum samples of individuals with convalescence Oropouche virus infection**

Blood samples from individuals with previous Oropouche virus (OROV) infection were obtained by venipuncture in May 2016 from residents of Coari municipality, Amazonas State, Brazil. The previous OROV infection was confirmed by plaque reduction neutralization test value 50 ($PRNT_{50}$) [1]. All the samples used in this study were negative by real-time quantitative Reverse Transcription-Polymerase Chain Reaction (RT-qPCR) for OROV, Mayaro (MAYV), chikungunya (CHIKV), and dengue viruses (DENV), as described (Table S5, p. 7). All samples were stored at –80°C.

**Real-time Quantitative Reverse Transcription-Polymerase Chain Reaction for Oropouche, *Chikungunya, Dengue, and Mayaro Viruses***

Viral RNA was extracted from the serum samples using the Maxwell HT Viral TNA Kit (Cat no. AX2340, Promega, USA) with the KingFisher Flex Purification System robot (Thermo Fisher Scientific, USA), following the manufacturer's instructions. The extracted RNA was then tested by real-time RT-qPCR targeting OROV[2,3], CHIKV[4], DENV serotypes 1 to 4[5,6], and MAYV[7] using the qPCR BIO Probe 1-Step Go Lo - ROX Kit (Cat no. PB25.11-03, PCR Biosystems, UK). Reactions were performed on QuantStudio 3 (Applied Biosystems, USA). The primers and probes used for the viral detection are described (Table S5, p. 7).

**Oropouche virus isolation in cell culture**

OROV isolation in culture cells was performed by inoculating Vero CCL-81 cells with ten serum samples that tested positive for OROV RNA using RT-qPCR (Table S1, p. 4). Briefly, Vero CCL-81 cells were plated in 24-well plates at a concentration of $2{\cdot}5{\times}10^5$ cells per mL ($1{\cdot}25x10^5$ cells per well) in Minimum Essential Eagle's Medium (DMEM) supplemented with 10% fetal bovine serum (FBS), and 1% of penicillin of 10,000 units and 10,000 µg/mL streptomycin solution. Subsequently, the serum samples (n=10) were diluted 1:10 in DMEM, treated with 2% penicillin and streptomycin, and added to the monolayer. After a one-hour incubation at 37ºC for adsorption, DMEM supplemented with 5% FBS and treated with 1% penicillin and streptomycin was added to the monolayer for maintenance. The cells were kept at 37ºC with 5% $CO_2$ and monitored for 30 hours until the cytopathic effect (CPE) became visible on an optical microscope. Next, the supernatant was collected and subjected to an RT-qPCR assay[3] to confirm viral isolation, indicated by a decrease in the Ct-value.

**Focus forming assay for Oropouche virus**

A focus formation assay was performed for the titration of OROV-positive serum samples and isolates, as previously described elsewhere[8]. Briefly, the samples were serially diluted in an 8-fold series in DMEM treated with 1% penicillin and streptomycin solution. Next, 100 µL of the dilutions were transferred to 96-well plates containing Vero CCL-81 cells ($5{\times}10^4$ cells per well) with 80% confluence, which were incubated for 1 hour at 37°C with 5% $CO_2$ for viral adsorption. Subsequently, 125 µL of DMEM containing 0·75% carboxymethylcellulose and 5% FBS was added to the wells, and the plates were incubated at 37°C with 5% $CO_2$ for 48 hours. Next, the cells were fixed with 70 µL of 8% paraformaldehyde solution (PFA) and incubated for 1 hour at room temperature. After removing the PFA, the cells were washed with phosphate-buffered saline (PBS). The cell monolayer was then blocked for 30 minutes with 150 µL of blotto. After blocking, the monolayers were washed with Perm/Wash Buffer (PBS supplemented with 0·1% BSA and 0·1% Triton X-100) and incubated with the polyclonal anti-OROV antibody (Cat no. VR-1228AF,

ATTC, USA). After a second wash with PermWash, the monolayers were incubated with an anti-mouse IgG secondary antibody (Cat no. AP124P, Sigma-Aldrich, USA). Finally, after a final wash with Perm/Wash Buffer, the assay was revealed using the True-Blue Peroxidase substrate (Cat no. 5510-0030, KPL, USA) for 30 minutes.

**Oropouche virus genome sequencing and analysis**
OROV genome sequencing was performed with two viral isolates using the Rapid SMART-9N protocol with the MinION platform (Oxford Nanopore Technologies, UK), as previously described[9]. The generated raw FAST5 files were then basecalled, demultiplexed, and trimmed using Guppy version 9·4·1 (Oxford Nanopore Technologies, UK). The barcoded files were aligned to the OROV reference genome (GenBank accession no. KP691612, KP691622, and KP691623) using minimap2 v·2·17·r941[10] and converted into BAM files using SAMtools[11]. Medaka_variants were employed for variant calling, followed by medaka_consensus (Oxford Nanopore Technologies, UK) for consensus sequence building. Genome regions with coverage below 20× were represented by the letter "N". NanoStat version 1·6·0[12], Samtools stats, and Samtools depth [12] were applied to compute the genome statistics.

**Phylogenetic analysis**
The two novel OROV genomes with >90% coverage were generated and aligned with the non-redundant OROV strains with complete coding sequences available in the GenBank database as of June 30, 2024. Then, we built a multiple sequence alignment (MSA) for each segment using MAFFT version 7·450[13], and manual adjustment was conducted using Geneious Prime 2023·0·4. A maximum likelihood (ML) phylogeny trees were performed using IQ-TREE version 2 under a GTR+I+γ model determined by ModelFinder[14,15]. The ultrafast-bootstrap approach with 1,000 replicates was used to determine the statistical support for nodes for the ML phylogenies. The phylogenetic trees were visualized using Figtree 1·4·2 (http://tree.bio.ed.ac.uk/software/figtree/). Additionally, we contacted the three segments of genomes, and we screened for reassortment events using all available methods in RDP version 4[16].

**Plaque reduction neutralization test for Oropouche virus**
To compare the neutralizing antibody capacity of serum from individuals previously infected with OROV against OROV strains BeAn 19991 (prototype) and AM0088 strain (2023-2024 OROV reassortment), we performed a $PRNT_{50}$ as described elsewhere[1]. Briefly, we inactivated the complement system by heating serum samples at 56ºC degrees, then we performed serial dilutions of each serum sample and incubated with a solution containing $2\times10^3$ PFU/mL for BeAn 19991 isolate, or 80 PFU/mL of the AM0088 isolate, both for 1 hour at 37°C. Subsequently, the virus-serum mixtures were added to pre-formed Vero CCL-81 cell monolayers and incubated for 1 hour at 37°C in a 5% $CO_2$ atmosphere. Next, we removed the inoculum and added 1 mL of DMEM containing 0·75% carboxymethylcellulose and 5% FBS was gently added to each well, and the plates were incubated at 37°C in a 5% $CO_2$ atmosphere for 3 days. Finally, the cells were fixed with 500 µL of 8% paraformaldehyde solution for 1 hour and stained with 1% methylene blue (Cat no. PHR3838, Sigma-Aldrich, USA) for 5 minutes. Plaque reduction was calculated as the average of values from two technical duplicates, corresponding to the percentages of the number of plaques counted compared to the positive control. These values were transformed to $Log_2$ for better visualization in the graph and subjected to a three-parameter nonlinear dose-response inhibition regression test.

**Virus replication curves for Oropouche virus**

To compare the viral fitness OROV strains BeAn 19991 (prototype) or AM0088 (2023-2024 OROV reassortment), we performed virus replication curves using Vero CCL-81 cells (African green monkey kidney), Huh7 cells (human liver carcinoma), and U-251 cells (Human glioblastoma astrocytoma). In summary, the cells were infected with OROV strains BeAn 19991 or AM0088 at a MOI of 0·1 for 1 hour at 37°C in a 5% $CO_2$ atmosphere. Then, we removed the inoculum, washed the cell monolayer three times using PBS, and added Minimum Essential Eagle's Medium (DMEM) supplemented with 10% fetal bovine serum (FBS), and 1% of penicillin of 10,000 units and 10,000 µg/mL streptomycin solution. At 3-, 6-, 12-, and 24-hours post-infection (hpi), we collected the cell culture supernatant and determined the infectious virus using FFA as described above[8]. All the experiments were conducted in triplicate.

#### Assessment of plaque phenotypes of Oropouche virus

To evaluate the plaque phenotypes generated by OROV strains BeAn 19991 (prototype) or AM0088 (2023-2024 OROV reassortment), we counted the number of plaques and measured the size of plaques at 36-, 48-, and 72-hours post-infection (hpi) for both OROV isolate in Vero CCL-81 cells. The number of plaques produced by each strain at each time point was determined by visual observation and counting. To measure the plaques, we illuminated the plates with a white bottom light and took photos using a Canon EOS Rebel T7i at a focal distance of 54 mm. All assay images were then imported into Fiji software version 2·15·1 for further analysis. We pre-processed and filtered the images to obtain a clearer resolution of the plaques. Next, we used the "Analyze Particles" feature to identify and measure the plaques, using the well diameter as the reference scale. Finally, any identified noise was manually removed from the images.

#### Cross-neutralization antibody test for Oropouche virus

To investigate antigenic differences between the OROV BeAn 19991 and AM0088 isolates, we conducted cross-neutralization assays. Briefly, two groups of four-week-old C57BL/6 mice were intraperitoneally (IP) inoculated with $1\times10^6$ PFU using a final volume of 100 µl. One group was inoculated with the BeAn 19991 isolate, and the other with AM0088 isolate. The mice were then kept under pathogen-free conditions at the biosafety level 2 animal facility of the Institute of Biology at the University of Campinas. Next, serum from these animals was collected 28 days post-infection. All animals did not show any signs of disease. Then, we performed the $PRNT_{50}$ to evaluate the capacity of neutralizing antibodies against the same OROV isolate (homologous) or different OROV isolate (heterologous).

#### Supplementary Figure

**Fig. S1.** Mismatches between the segment S of 2023-2024 OROV reassortment strains and the primers and probes previously described[2].

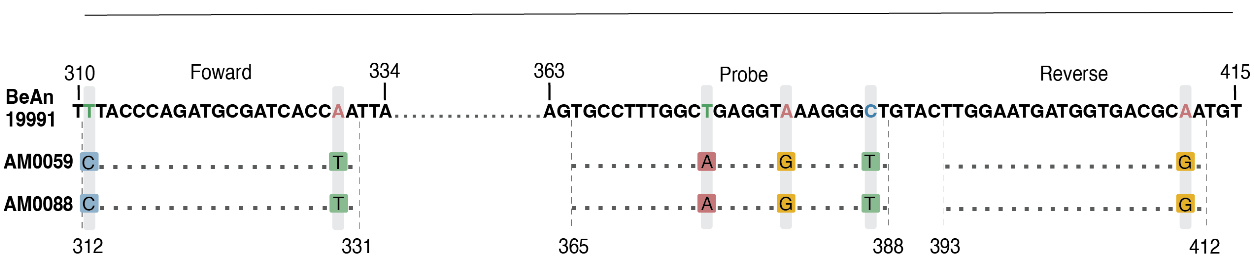

### Supplementary Table

**Table S1.** Anonymized patient information data for all PCR-positive samples of Oropouche virus.

| Group | ID | Sample collection | RT-PCR-SegS[3] | RT-PCR-SegM[3] | RT-PCR-SegS[2] | Viral isolate | Sequencing |
|---|---|---|---|---|---|---|---|
| Febrile illnesses | 0004 | 12-Jan-2024 | 36·4 | 32·1 | ND | yes | N/P |
| | 0044 | 16-Jan-2024 | 37·0 | ND | ND | no | N/P |
| | 0045 | 19-Jan-2024 | 37·3 | 29·7 | ND | yes | N/P |
| | 0046 | 21-Jan-2024 | 35·2 | 28·0 | ND | yes | N/P |
| | 0059 | 21-Jan-2024 | 34·5 | 28·6 | ND | yes | yes |
| | 0063 | 20-Jan-2024 | 36·9 | 35·0 | ND | yes | N/P |
| | 0064 | 21-Jan-2024 | 36·5 | 30·0 | ND | yes | N/P |
| | 0078 | 28-Jan-2024 | 34·2 | 26·0 | ND | no | N/P |
| | 0088 | 30-Jan-2024 | 36·1 | 28·7 | ND | yes | yes |
| | 0093 | 22-Jan-2024 | 36·3 | not detected | ND | no | N/P |
| CNS manifestations | 2051 | 20-Jan-2024 | N/P | 32·8 | ND | N/P | N/P |
| | 2062 | 31-Jan-2024 | N/P | 33·9 | ND | N/P | N/P |
| | 2134 | 16-Apr-24 | N/P | 36·9 | ND | N/P | N/P |

Seg S. segment S. SegM, segment M. CNS, central nervous system. N/P, not performed. ND, not detected.

**Table S2.** Amino acids change comparing OROV strain BeAn19991 to 2023-2024 OROV reassortment strains (AM0059 and AM0088).

| Gene | Position | BeAn19991 strain | AM0059 strain | AM0088 strain |
|---|---|---|---|---|
| M | 12 | G | S | S |
| | 22 | S | S | N |
| | 66 | K | R | R |
| | 71 | del | T | T |
| | 72 | del | T | T |
| | 247 | F | F | S |
| | 276 | F | I | I |
| | 394 | R | S | S |
| | 395 | I | T | T |
| | 430 | D | N | N |
| | 448 | V | I | I |
| | 516 | S | N | N |
| | 523 | F | S | S |
| | 616 | N | K | K |
| | 645 | M | I | I |
| | 734 | L | V | V |
| | 752 | G | D | D |
| | 766 | N | S | S |
| | 814 | S | S | P |
| | 824 | A | T | T |
| | 848 | I | I | V |
| | 959 | I | I | V |
| | 983 | Q | K | K |
| | 984 | S | G | G |
| | 1085 | V | A | A |
| | 1205 | I | T | T |
| | 1270 | P | S | S |
| | 1311 | T | I | I |
| | 1325 | I | V | V |
| | 1344 | R | K | K |
| | 1362 | I | V | V |
| L | 135 | T | A | A |
| | 144 | V | M | M |
| | 201 | T | A | A |

| | | | |
|---|---|---|---|
| 210 | S | N | N |
| 215 | A | S | S |
| 258 | H | Q | Q |
| 263 | T | A | A |
| 284 | N | S | S |
| 303 | M | I | I |
| 309 | K | Q | Q |
| 313 | N | S | S |
| 338 | V | I | I |
| 339 | N | S | S |
| 354 | V | I | I |
| 372 | I | V | V |
| 382 | I | V | V |
| 415 | L | F | F |
| 442 | N | D | D |
| 458 | I | T | T |
| 464 | I | V | V |
| 558 | M | I | I |
| 565 | A | T | T |
| 580 | A | T | T |
| 663 | R | K | K |
| 677 | S | A | A |
| 786 | A | V | A |
| 788 | R | Q | Q |
| 789 | L | T | T |
| 790 | S | V | V |
| 791 | X | N | N |
| 794 | V | I | I |
| 799 | L | I | I |
| 800 | Q | A | A |
| 801 | E | R | R |
| 802 | X | N | N |
| 850 | R | K | K |
| 853 | L | T | T |
| 854 | R | K | K |
| 855 | M | N | N |
| 856 | I | D | D |
| 857 | Q | A | A |
| 921 | N | S | S |
| 940 | H | Y | Y |
| 1035 | N | S | S |
| 1114 | L | V | V |
| 1159 | I | T | T |
| 1192 | I | V | V |
| 1314 | S | N | N |
| 1375 | K | R | R |
| 1436 | D | N | N |
| 1439 | A | T | T |
| 1505 | V | I | I |
| 1693 | V | I | I |
| 1758 | V | I | I |
| 1778 | I | V | V |
| 1911 | R | Q | Q |
| 1934 | V | I | I |
| 1942 | V | I | I |
| 1948 | S | N | N |
| 1961 | I | V | V |
| 1976 | R | K | K |
| 2056 | G | E | E |
| 2057 | D | N | N |
| 2171 | K | R | R |
| 2187 | I | V | V |

The positions refer to the competing coding sequences of OROV strain BeAn 19991 for the M segment (GenBank accession number NC_005775) and the L segment (GenBank accession number NC_005776).

**Table S3.** Neutralizing capacity of serum previously infected with OROV in Coari municipality, Amazonas State, Brazil against the BeAn 19991 and the AM0088 isolates.

| ID | BeAn 19991 | AM0088 |
|---|---|---|
| A 19 | 640 | <20 |
| A 20 | 640 | <20 |
| A 24 | 160 | <20 |
| A 33 | 640 | <20 |
| A 35 | 160 | <20 |
| A 77 | 320 | <20 |
| A 91 | 640 | <20 |
| A 111 | 320 | <20 |
| A 193 | 640 | <20 |
| A 206 | 640 | <20 |
| A 216 | 640 | <20 |
| A 304 | 320 | <20 |
| A 324 | 640 | <20 |
| A 336 | 640 | <20 |
| A 340 | 320 | <20 |
| A 351 | 640 | <20 |
| A 372 | 160 | <20 |
| A 375 | 320 | <20 |
| A 385 | 640 | <20 |
| A 389 | 320 | <20 |
| A 392 | 320 | <20 |
| A 369 | 640 | <20 |

**Table S4.** Genome sequences used in the phylogenetic analyses.

| Isolate | Country | State | Host | Collection year | Accession GenBank numbers | | |
|---|---|---|---|---|---|---|---|
| TRVL-9760 | Trinidad and Tobago | - | Homo sapiens | 1955 | KC759124 | KC759123 | KC759122 |
| BeH759620 | Brazil | Amapá | Homo sapiens | 2009 | KP691623 | KP691622 | KP691621 |
| TVP-19250/GML-444839 | Panama | - | Homo sapiens | 1989 | KP795080 | KP795079 | KP795078 |
| TVP-19256/IQE-7894 | Peru | - | Homo sapiens | 2008 | KP795086 | KP795085 | KP795084 |
| TVP-19257/IQT-1690 | Peru | - | Homo sapiens | 1995 | KP795089 | KP795088 | KP795087 |
| TVP-19258/IQT-4083 | Peru | - | Homo sapiens | 1997 | KP795092 | KP795091 | KP795090 |
| TVP-19259/IQT-7085 | Peru | - | Homo sapiens | 1998 | KP795095 | KP795094 | KP795093 |
| TVP-19260/MD-203 | Peru | - | Homo sapiens | 1994 | KP795098 | KP795097 | KP795096 |
| TVP-19264/PAN-481126 | Panama | - | Homo sapiens | 1999 | KP795104 | KP795103 | KP795102 |
| 087/2016 | Ecuador | - | Homo sapiens | 2016 | MF926352 | MF926353 | MF926354 |
| BeH 389865 | Brazil | Amazonas | Homo sapiens | 1980 | MG747506 | MG747507 | MG747508 |
| BeH 390242 | Brazil | Amazonas | Homo sapiens | 1980 | MG747509 | MG747510 | MG747511 |
| BeH 472433 | Brazil | Maranhão | Homo sapiens | 1988 | MG747512 | MG747513 | MG747514 |
| BeH 472435 | Brazil | Maranhão | Homo sapiens | 1988 | MG747515 | MG747516 | MG747517 |
| BeH 421086 | Brazil | Maranhão | Homo sapiens | 1993 | MG747518 | MG747519 | MG747520 |
| BeAn 626990 | Brazil | Minas Gerais | Callithrix sp. | 2000 | MG747521 | MG747522 | MG747523 |
| BeAr 19886 | Brazil | Pará | Ochlerotatus serratus | 1960 | MG747524 | MG747525 | MG747526 |
| BeH 29086 | Brazil | Pará | Homo sapiens | 1961 | MG747527 | MG747528 | MG747529 |
| BeH 121923 | Brazil | Pará | Homo sapiens | 1967 | MG747533 | MG747534 | MG747535 |
| BeAr 136921 | Brazil | Pará | Culex quinquefasciatus | 1968 | MG747536 | MG747537 | MG747538 |
| BeAn 206119 | Brazil | Pará | Bradypus tridactylus | 1971 | MG747539 | MG747540 | MG747541 |
| BeH 355173 | Brazil | Pará | Homo sapiens | 1978 | MG747548 | MG747549 | MG747550 |
| BeAr 366927 | Brazil | Pará | Culicoides paraensis | 1979 | MG747551 | MG747552 | MG747553 |

| | | | | | | | |
|---|---|---|---|---|---|---|---|
| BeH 385591 | Brazil | Pará | Homo sapiens | 1980 | MG747554 | MG747555 | MG747556 |
| BeH 532314 | Brazil | Pará | Homo sapiens | 1994 | MG747557 | MG747558 | MG747559 |
| BeH 532490 | Brazil | Pará | Homo sapiens | 1994 | MG747563 | MG747564 | MG747565 |
| BeH 541140 | Brazil | Pará | Homo sapiens | 1994 | MG747569 | MG747570 | MG747571 |
| BeH 543857 | Brazil | Pará | Homo sapiens | 1996 | MG747578 | MG747579 | MG747580 |
| PPS 522 H 669314 | Brazil | Pará | Homo sapiens | 2003 | MG747581 | MG747582 | MG747583 |
| PMOH 682426 | Brazil | Pará | Homo sapiens | 2004 | MG747587 | MG747588 | MG747589 |
| PMOH 682431 | Brazil | Pará | Homo sapiens | 2004 | MG747590 | MG747591 | MG747592 |
| BeH 498913 | Brazil | Rondonia | Homo sapiens | 1990 | MG747602 | MG747603 | MG747604 |
| BeH 505768 | Brazil | Rondonia | Homo sapiens | 1991 | MG747605 | MG747606 | MG747607 |
| 057/2016 | Ecuador | - | Homo sapiens | 2016 | MK506818 | MK506823 | MK506828 |
| Haiti-1/2014 | Haiti | - | Homo sapiens | 2014 | MN264269 | MN264268 | MN264267 |
| Bel90435/H853382 | Brazil | Pará | Homo sapiens | 2018 | MT879230 | MT879229 | MT879228 |
| ZDC388 | Brazil | Rondonia | Homo sapiens | 2023 | PP153945 | PP153946 | PP153947 |
| 0628MJG | Brazil | Roraima | Homo sapiens | 2022 | PP153981 | PP153982 | PP153983 |
| 0026TSS | Brazil | Roraima | Homo sapiens | 2023 | PP154011 | PP154012 | PP154013 |
| 3896ERA | Brazil | Amazonas | Homo sapiens | 2023 | PP154038 | PP154042 | PP154043 |
| 0545 | Brazil | Acre | Homo sapiens | 2023 | PP154152 | PP154153 | PP154154 |
| ILMD_TF29 | Brazil | Amazonas | Homo sapiens | 2015 | PP154170 | PP154171 | PP154172 |
| BeH505764 | Brazil | Rondonia | Homo sapiens | 1991 | PP357049 | PP357049 | PP357048 |
| BeAn 19991 | Brazil | Pará | Bradypus tridactylus | 1955 | KP052852 | KP052851 | KP052850 |
| BeH 532314 | Brazil | Pará | Homo sapiens | 1994 | MG747559 | MG747558 | MG747557 |
| BeH 498913 | Brazil | Rondonia | Homo sapiens | 1990 | MG747602 | MG747603 | MG747604 |
| BeH 390242 | Brazil | Amapá | Homo sapiens | 1980 | MG747509 | MG747510 | MG74751 |
| BeH 385591 | Brazil | Pará | Homo sapiens | | MG747554 | MG747555 | MG747556 |
| BeAr 19886 | Brazil | Pará | Ochlerotatus serratus | 1960 | MG747524 | MG7475215 | MG747526 |
| BeAn 423380 | Brazil | Pará | Nasua nasua | 1984 | NC_043576 | NC_043577 | NC_043578 |

**Table S5.** Primers and probes used for the detection of arboviruses in this study.

| Virus | Sequences (5'→3') | Primers and probes | Target | Genome position | Ref. |
|---|---|---|---|---|---|
| OROV | TCCGGAGGCAGCATATGTG | Forward | S | 98-116 | [3] |
| | ACAACACCAGCATTGAGCACTT | Reverse | S | 160-139 | |
| | ATTTGAAGCTAGATACGG | Probe | | 118-136 | |
| | TACCCAGATGCGATCACCAA | Forward | S | 356-375 | [2] |
| | TTGCGTCACCATCATTCCAA | Reverse | S | 437-456 | |
| | TGCCTTTGGCTGAGGTAAAGGGCT | Probe | | 409-433 | |
| | TCGTCAACAAAACTCAACCACTTT | Forward | M | 428-451 | [2] |
| | GACCACAATTTACGGTTACATGCT | Reverse | M | 525-548 | |
| | TCGGGACAACTTCGACATCAGGCTG | Probe | | 459-483 | |
| DENV1 | GACACCACACCCTTTGGACAA | Forward | NS5 | 8586-8606 | [6] |
| | CACCTGGGCTGTCACCTCCAT | Reverse | NS5 | 8692-8673 | |
| | AGAGGGTGTTTAAAGAGAAAGTTGACACGCG | Probe | | 8608-8638 | |
| DENV2 | CAGGTTATGGCACTGTCACGAT | Forward | M | 1605 | [5] |
| | CCATCTGCAGCAACACCATCTC | Reverse | M | 1583 | |
| | CTCCGAGAACAGGCCTCGACTTCAA | Probe | | 1008 | |
| DENV3 | GGGAAAACCGTCTATCAATA | Forward | C | 118-221 | [6] |
| | CGCCATAACCAATTTCATTGG | Reverse | C | 241-221 | |
| | CACAGTTGGCGAAGAGATCTCAAGAGGA | Probe | | 174-202 | |
| DENV4 | TGAAGAGATTCTCAACCGGAC | Forward | C | 187-207 | [6] |
| | AATCCCTGCTGTTGGTGGC | Reverse | C | 293-275 | |

| | | | | | |
|---|---|---|---|---|---|
| | TCATCACGTTTTTGCGAGTCCTTTCCA | Probe | | 247-273 | |
| CHIKV | AAAGGGCAAACTCAGCTTCAC | Forward | NSP1 | 874-894 | 4 |
| | GCCTGGGCTCATCGTTATTC | Reverse | NSP1 | 961-942 | |
| | CGCTGTGATACAGTGGTTTCGTGTG | Probe | | 899-923 | |
| MAYV | AAGCTCTTCCTCTGCATTGC | Forward | NSP1 | 51-70 | 7 |
| | TGCTGGAAACGCTCTCTGTA | Reverse1 | NSP1 | 141-160 | |
| | TGCTGGAAATGCTCTTTGTA | Reverse2 | | 141-160 | |
| | GCCGAGAGCCCGTTTTTAAAATCA | Probe | | 116-140 | |

Legend: OROV, Oropouche virus. DENV, dengue virus. CHIKV, chikungunya virus. MAYV, Mayaro virus. S, S segment. M, M segment. NS5, non-structural protein 5. M, matrix protein. C, capsid protein. NSP1, non-structural protein 1.

### References


1. Proenca-Modena JL, Hyde JL, Sesti-Costa R, et al. Interferon-Regulatory Factor 5-Dependent Signaling Restricts Orthobunyavirus Dissemination to the Central Nervous System. *J Virol* 2016; **90**(1): 189-205.
2. de Souza Luna LK, Rodrigues AH, Santos RI, et al. Oropouche virus is detected in peripheral blood leukocytes from patients. *J Med Virol* 2017; **89**(6): 1108-11.
3. Naveca FG, Nascimento VAD, Souza VC, Nunes BTD, Rodrigues DSG, Vasconcelos PFDC. Multiplexed reverse transcription real-time polymerase chain reaction for simultaneous detection of Mayaro, Oropouche, and Oropouche-like viruses. *Mem Inst Oswaldo Cruz* 2017; **112**(7): 510-3.
4. Lanciotti RS, Kosoy OL, Laven JJ, et al. Chikungunya virus in US travelers returning from India, 2006. *Emerg Infect Dis* 2007; **13**(5): 764-7.
5. Johnson BW, Russell BJ, Lanciotti RS. Serotype-specific detection of dengue viruses in a fourplex real-time reverse transcriptase PCR assay. *J Clin Microbiol* 2005; **43**(10): 4977-83.
6. Callahan JD, Wu SJ, Dion-Schultz A, et al. Development and evaluation of serotype- and group-specific fluorogenic reverse transcriptase PCR (TaqMan) assays for dengue virus. *J Clin Microbiol* 2001; **39**(11): 4119-24.
7. Waggoner JJ, Rojas A, Mohamed-Hadley A, de Guillén YA, Pinsky BA. Real-time RT-PCR for Mayaro virus detection in plasma and urine. *J Clin Virol* 2018; **98**: 1-4.
8. Proenca-Modena JL, Sesti-Costa R, Pinto AK, et al. Oropouche virus infection and pathogenesis are restricted by MAVS, IRF-3, IRF-7, and type I interferon signaling pathways in nonmyeloid cells. *J Virol* 2015; **89**(9): 4720-37.
9. Claro IM, Ramundo MS, Coletti TM, et al. Rapid viral metagenomics using SMART-9N amplification and nanopore sequencing. *Wellcome Open Res* 2021; **6**: 241.
10. Li H. Minimap2: pairwise alignment for nucleotide sequences. *Bioinformatics* 2018; **34**(18): 3094-100.
11. Li H, Handsaker B, Wysoker A, et al. The Sequence Alignment/Map format and SAMtools. *Bioinformatics* 2009; **25**(16): 2078-9.
12. De Coster W, D'Hert S, Schultz DT, Cruts M, Van Broeckhoven C. NanoPack: visualizing and processing long-read sequencing data. *Bioinformatics* 2018; **34**(15): 2666-9.
13. Katoh K, Standley DM. MAFFT multiple sequence alignment software version 7: improvements in performance and usability. *Mol Biol Evol* 2013; **30**(4): 772-80.
14. Nguyen LT, Schmidt HA, von Haeseler A, Minh BQ. IQ-TREE: a fast and effective stochastic algorithm for estimating maximum-likelihood phylogenies. *Mol Biol Evol* 2015; **32**(1): 268-74.

15. Kalyaanamoorthy S, Minh BQ, Wong TKF, von Haeseler A, Jermiin LS. ModelFinder: fast model selection for accurate phylogenetic estimates. *Nat Methods* 2017; **14**(6): 587-9.
16. Martin DP, Murrell B, Golden M, Khoosal A, Muhire B. RDP4: Detection and analysis of recombination patterns in virus genomes. *Virus Evol* 2015; **1**(1): vev003.
